# The Day-1 Mobility Loss Model: Development, Validation, and Clinical Applicability of a Model to Predict Hospital Acquired Mobility Loss in Older Adults

**DOI:** 10.1101/2022.08.22.22279075

**Authors:** Sachin J. Shah, Ari Hoffman, Logan Pierce, Kenneth E. Covinsky

**Author notes:** <u>Corresponding author</u> Sachin J Shah, MD, MPH, 100 Cambridge St, Suite 1600, Boston, MA 02114, @sachinjshah.

## Abstract

**Background:** Mobility loss is common in hospitalized older adults, and resources to prevent mobility loss are finite. Our goal was to develop a rapid, universal screening tool that identifies individuals at risk of losing the ability to walk during hospitalization on the first hospital day. Second, we determined if the model could inform the use of mobility-preserving interventions.

**Methods:** We included patients admitted to a general medical service, aged ≥65 years, who could walk on admission (Braden Scale Activity subset >=3). Patients were considered to have a new mobility impairment if the activity score was <3 on discharge. We used predictors available on the first hospital day to develop (2017-18) and validate (2019) a prediction model. We determined the association between predicted risk and therapy use in the validation cohort to highlight the model’s clinical utility.

**Results:** 5542 patients were included (median age 76yrs, 48% women); 7.6% were discharged unable to walk. The model included six predictors: age, marital status, medication administrations, Glasgow Coma Scale verbal score, serum albumin, and urinary catheter presence. In the validation cohort, the model discriminated well (c-statistic 0.75) and was strongly associated with hospital-acquired mobility loss (lowest decile 1%, highest decile 24%). In the validation cohort, therapy consultation ordering increased linearly with predicted risk; however, observed mobility loss increased exponentially.

**Conclusion:** The Day-1 Mobility Loss model predicts the risk of mobility loss in hospitalized older adults on the first hospital day. Further, it identifies at-risk older adults who may benefit from mobility interventions.

## INTRODUCTION

Often overlooked, mobility is vital to the care of the hospitalized older adult. Immobility is associated with falls, pressure injuries, delirium, and discharge to a rehabilitation facility.^1–5^ Yet immobility remains highly prevalent in U.S. hospitals—most patients spend less than 3% of the day standing or walking.^6,7^ Older adults are particularly susceptible to the ill effects of immobility; one in three patients over the age of 65 loses the ability to independently perform one or more activities of daily living following hospital admission.^8,9^ Worse yet, functional impairment can persist well after discharge.^10,11^

While programs to improve mobility are promising, targeting and implementation remain hurdles to widespread adoption. Mobility programs have been shown to prevent loss of function in inpatient settings, from the intensive care unit to the general medical floor to those admitted for arthroplasty.^12–15^ While effective, such programs are challenging to implement in hospitals because resources are finite—there are too few therapists, ACE unit beds, and mobility program staff for all hospitalized older adults. In this setting, predicting which patients are most likely to develop hospital-acquired disability enables targeting of finite resources.

Multiple screening instruments and models have been developed to predict inpatient loss of function; however, none can reasonably support universal screening.^16–21^ Existing instruments use demographics, social characteristics, clinical features (e.g., lab values, comorbidities), and functional assessments (e.g., cognitive assessment using the Mini-Mental Status Exam). While useful, the current landscape lacks a screening tool that relies solely on routinely collected electronic medical record (EMR) data and does not require added clinician input. Such a tool could support universal screening. To address this gap, we addressed two objectives. First, we developed the Day-1 Mobility Loss Model, a model to predict mobility loss using available data in the EMR in the first 24 hours of hospital admission. Second, we determined if the model could meaningfully inform clinicians’ use of therapy consultations.

## METHODS

### Study cohort and Data

We examined all patients admitted to the Hospital Medicine Service at the University of California San Francisco Hospital over three years before the COVID19 pandemic (Jan 1, 2017, through Dec 31, 2019). We included adults aged ≥65 years who were documented by their bedside nurse to be walking on their first hospital day. Mobility was assessed using the activity subscale of the Braden Scale for Predicting Pressure Sore Risk (Braden Scale).^22^ The Braden activity subscale categorizes patients into four mobility levels and associated scores: Bedfast (score = 1), Chairfast (score = 2), Walks occasionally (score = 3), Walks frequently (score = 4) (**Appendix 1**). At the study site, bedside nurses measured the Braden score two to three times per day which we averaged to obtain a daily score. Patients were included and considered to have no mobility impairment if their mean score was ≥3 in the first 24 hours of admission. Predictors were obtained using administrative and electronic medical record data, including nursing evaluation data. We excluded patients with missing outcome data (<1%) (**Appendix 2**).

### Outcome Measurement

We defined new-onset hospital-acquired mobility impairment as a change in the Braden Score activity subscale from >=3 on admission to <3 in the 24 hours before discharge. This definition build on prior work using the Braden activity subscale to study mobility in hospitalized patients.^23^

### Predictors

We sourced predictors using the conceptual model for hospital-acquired mobility impairment by Chase et al.^24^ that were also available on the first hospital day. Specifically, we identified demographic factors (age, marital status, self-reported race and gender, insurance status), clinical factors (transfer admission, unique inpatient medication count and count of medications administered in first 24 hours, mental status using the Glasgow Coma Scale [GCS]^25,26^, serum creatinine, serum albumin, NPO status), and environmental factors (peripheral intravenous lines, gastric tube, urinary catheter). Medication administrations combine unique medications with frequency (e.g., vancomycin administered three times, give a value of 3) to reflect complexity and intensity of treatment. Missing predictors were handled individually. No patients were missing age, sex, insurance status, admit source, peripheral intravenous lines status, medication count, medication administrations, urinary catheter status, gastric tube status, or NPO status. Patients with missing, unknown, or other marital status and self-reported race were categorized together as a separate group, and these values were not imputed. When GCS scores, serum creatinine, or serum albumin were not recorded on the first hospital day, they were assumed to be normal (i.e., GCS 15, creatinine 1.0 mg/dL, albumin 4.0 gm/dL).

### Model development, validation, and test characteristics

We used calendar years 2017 and 2018 data to develop the prediction model. We determined the functional form of continuous variables by assessing linear, log, exponential, and clinical meaningful categorization against the outcome; we used the functional form with the lowest Bayesian Information Criterion (**Appendix 3**). We created 1000 bootstrapped samples with replacement of the development set. Using hospital-acquired mobility impairment as the outcome, we fit a logistic regression model using backward selection with a P value of <0.05 for a predictor to stay in the model. We then selected predictors that appeared in more than 60% of the bootstrap sample models. We then validated the model using calendar year 2019 data. We determined discrimination (c-statistic) and calibration (calibration slope and intercept) in the validation data.^27^ We and others have used this approach in prior studies.^28,29^

### Model application

We conducted two analyses to demonstrate the clinical utility of the prediction model. First, we sought to determine if the model could supplement clinical decision-making. To determine clinician perception of risk, we measured the association of therapy consultation order (Physical or Occupational therapy) on the first hospital day with predicted risk in the validation cohort. We inferred that, among other reasons, physicians worried about mobility decline would order a Physical or Occupational Therapy consultation. To assess if the model identifies patients at risk beyond clinical concern, we examined the association of predicted risk and observed mobility loss in patients who did not have a therapy consultation ordered on the first hospital day.

Second, we identified screening thresholds where this model could be used as a ‘rule-out’ test with high sensitivity (positive at 20^th^ percentile of risk) and a ‘rule-in’ test with high specificity (positive at 80^th^ percentile of risk). We report all results with 95% confidence intervals. We performed analyses using SAS 9.4 (Cary, NC) and R 4.0.2 (Vienna, Austria). The TRIPOD checklist can be found in **Appendix 6**. The University of California, San Francisco, Committee on Human Research approved the analyses for this study and waived the requirement for patient consent (Institutional Review Board No. 16-20781).

## RESULTS

### Patient characteristics

5542 patients met inclusion criteria. In the development cohort, the median age was 76 years (interquartile range [IQR], 69, 84), and 50% were women (**Table 1**). All patients were documented walking on admission, and 7.6% were discharged with a new mobility impairment. On the first hospital day, the median medication administrations was 29 (IQR 19, 42), 6% had a urinary catheter placed, and 33% were made NPO. Patient characteristics were similar when comparing the development and validation cohorts.

**Table 1:** Baseline patient characteristics of the derivation and validation cohorts.

|  | Development cohort<br>(n = 3570) | Validation cohort<br>(n = 1950) |
| --- | --- | --- |
| **Sociodemographic** |  |  |
| Age, median (IQR) | 76 (69, 84) | 75 (69, 83) |
| Marital status, % (no.) |  |  |
| Married | 50% (1778) | 51% (994) |
| Divorced | 9% (310) | 9% (178) |
| Widowed | 20% (708) | 17% (337) |
| Single | 21% (734) | 21% (417) |
| Other/Unknown/Declined | 1% (40) | 1% (24) |
| Race, % (no.) |  |  |
| White or Caucasian | 48% (1721) | 51% (1000) |
| Asian | 30% (1078) | 28% (537) |
| Black or African American | 9% (323) | 8% (160) |
| Other/Unknown/Declined | 13% (448) | 13% (253) |
| Patient gender, % (no.) |  |  |
| Men | 50% (1800) | 54% (1049) |
| Women | 50% (1770) | 46% (901) |
| Insurance status, % (no.) |  |  |
| Medicare | 73% (2620) | 72% (1406) |
| Medicare Advantage | 17% (600) | 17% (337) |
| Commercial | 5% (192) | 5% (103) |
| Medicaid | 4% (158) | 5% (104) |
| **Clinical** |  |  |
| Admit Source, % (no.) |  |  |
| Community | 97% (3462) | 96% (1871) |
| Transfer | 3% (108) | 4% (79) |
| Peripheral IV Count, median (IQR) | 1 (1, 2) | 1 (1, 2) |
| Unique medication count, median (IQR) | 7 (4, 10) | 7 (4, 10) |
| Medication administrations, median (IQR) | 29 (19, 42) | 29 (19, 42) |
| GCS Eyes, % (no.) |  |  |
| Normal (score = 4) | 97% (3468) | 97% (1890) |
| Abnormal (score < 4) | 3% (102) | 3% (60) |
| GCS Verbal, % (no.) |  |  |
| Normal (score = 5) | 90% (3570) | 88% (1708) |
| Abnormal (score < 5) | 10% (367) | 12% (242) |
| GCS Motor, % (no.) |  |  |
| Normal (score = 6) | 99% (3524) | 98% (1915) |
| Abnormal (score < 6) | 1% (46) | 2% (35) |
| Serum creatinine (mg/dL), median (IQR)* | 0.94 (0.72 - 1.34) | 0.95 (0.73 - 1.31) |
| Serum albumin (g/dL), median (IQR)** | 4.0 (3.3 - 4.0) | 4.0 (3.2 - 4.0) |
| Patient has urinary catheter, % (no.) |  |  |
| Yes | 6% (201) | 5% (99) |
| No | 94% (3369) | 95% (1851) |
| Patient has feeding tube, % (no.) |  |  |
| Yes | 2% (58) | 1% (14) |
| No | 98% (3512) | 99% (1936) |
| Patient made NPO, % (no.) |  |  |
| Yes | 35% (1255) | 38% (750) |
| No | 65% (2315) | 62% (1200) |

### Measures of model overall model performance

The final Day-1 Mobility Loss model included 6 variables to predict in-patient mobility loss: age, marital status, medication administration count, GCS verbal, albumin, urinary catheter placement (**Table 2**). Beyond age, which is a potent predictor of mobility loss, notable predictors include divorced marital status was also predictive (OR 1.78, 95% CI 1.14-2.76), medication administration count (19 vs. 42 administrations, OR 2.44, 95% CI 2.02 to 2.94), abnormal GCS verbal score (OR 2.60, 95%CI 1.86 to 3.62), serum albumin (3.3 g/dL vs. 4.0 g/dL, OR 1.34, 95% CI 1.27-1.42) and presence of a urinary catheter (OR 2.22, 95% CI 1.48 to 3.33). For representation and interpretability, odds ratios for log-transformed predictors (i.e., medication administrations, albumin) are listed for the 25th vs. 75th percentile.

**Table 2:** Predictors and odds ratios in final model used to predict hospital-acquired mobility loss.

| **Predictor** | **Odds ratio (95% CI)** |
| --- | --- |
| Patient age (84 vs. 69 yrs.) * | 1.94 (1.52-2.76) |
| Marital status |  |
| Married | 1.0 (ref) |
| Divorced | 1.78 (1.14 - 2.76) |
| Widowed | 1.06 (0.74 - 1.52) |
| Single | 1.36 (0.97 - 1.92) |
| Unknown/Declined | 2.61 (1.04 - 6.54) |
| Medication administrations (42 vs. 19 administrations) * | 2.44 (2.02 – 2.94) |
| GCS Verbal |  |
| Normal (GCS verbal = 5) | 1.0 (ref) |
| Abnormal (GCS verbal < 5) | 2.60 (1.86 - 3.62) |
| Serum albumin (3.3 vs 4.0 g/dL) * | 1.34 (1.27-1.42) |
| Urinary catheter |  |
| No | 1.0 (ref) |
| Yes | 2.22 (1.48 - 3.33) |

GCS—Glasgow coma scale.
\* For representation and interpretability odds ratios for continuous (age) and log transformed predictors (medication administrations, albumin) are listed for the 75<sup>th</sup> vs 25<sup>th</sup> percentile.

### Validation measures

The model performed well in the validation cohort. The model was well calibrated— observed and expected mobility loss in the validation cohort were highly correlated (R^2^ 0.97, calibration slope 0.93, **Appendix 4**). In the lowest decile, the observed mobility loss was 1.0% (predicted 1.3%), and the highest observed mobility loss was 23.6% (predicted 25.8%). The model discriminated well with an AUC of 0.75 (95% CI 0.72-0.78) to predict mobility loss in the development cohort and an AUC of 0.75 (95% CI 0.71 to 0.79) in the validation cohort (**Appendix 5**).

### Clinical utility

**Figure 2** and **Figure 3** together demonstrate that the Day 1 Mobility Loss model can supplement clinical decisions to target therapy resources. **Figure 2** shows the association between the predicted risk of mobility loss and therapy consultation order on the first hospital day in the validation cohort. The rate of therapy ordering on the first hospital day increased by an absolute 2.1% for every 5^th^ percentile increase in predicted risk, demonstrating a linear response to predicted risk.

**Figure 1:**
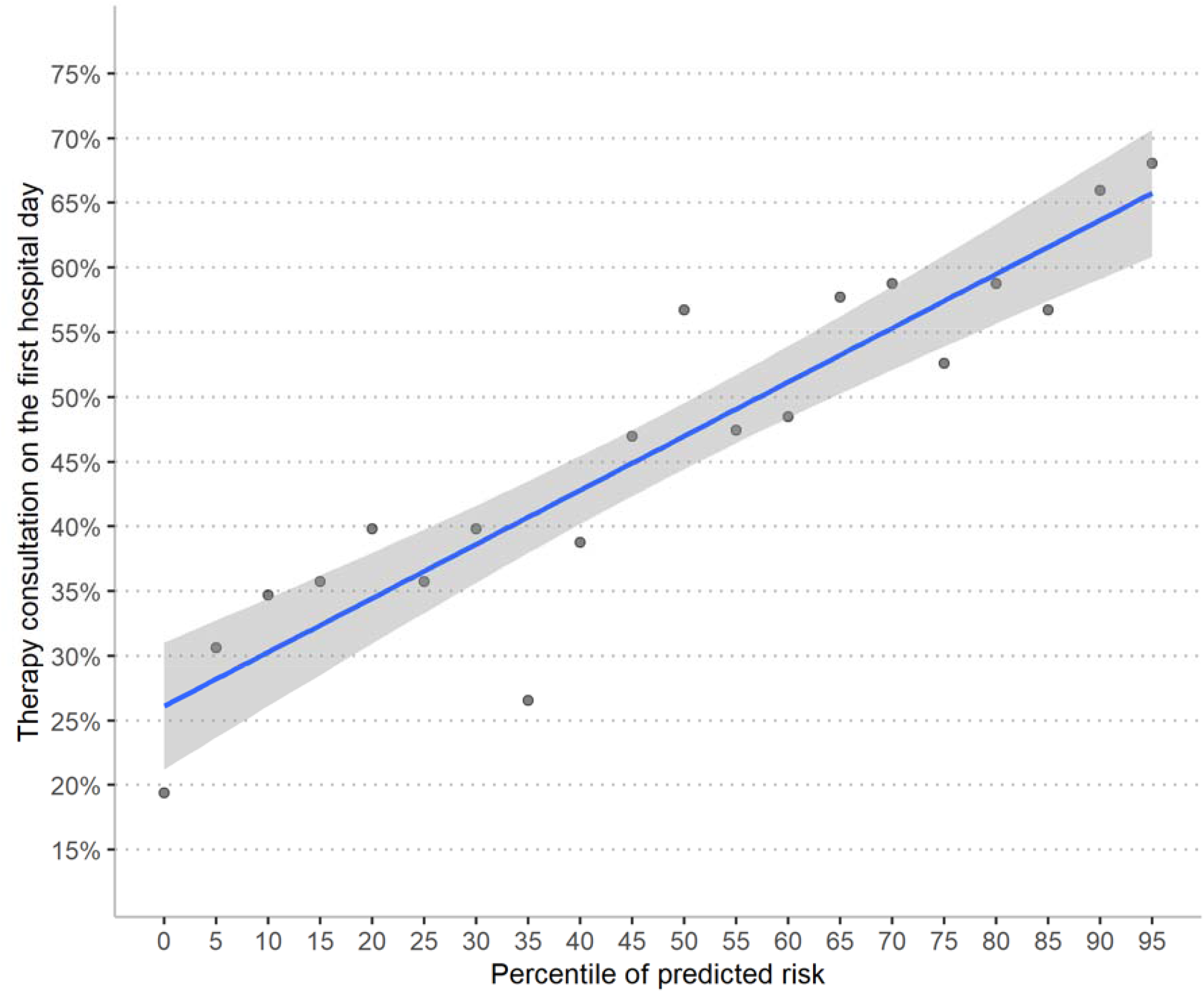
Physical or occupational therapy consultation ordering on the first hospital day by predicted risk, validation cohort. <u>Legend</u> Analysis performed in the validation data set. Graph displays therapy consultation order rate on the first hospital day by ventile of predicted risk. The best fit line is from a linear regression. The rate of therapy ordering increases by an absolute 2.1% (95% CI 1.7 to 2.5%) for every 5^th^ percentile increase in predicted risk. At the lowest predicted risk percentile (i.e., intercept) 24% of patients have a therapy consultation ordered.

**Figure 2:**
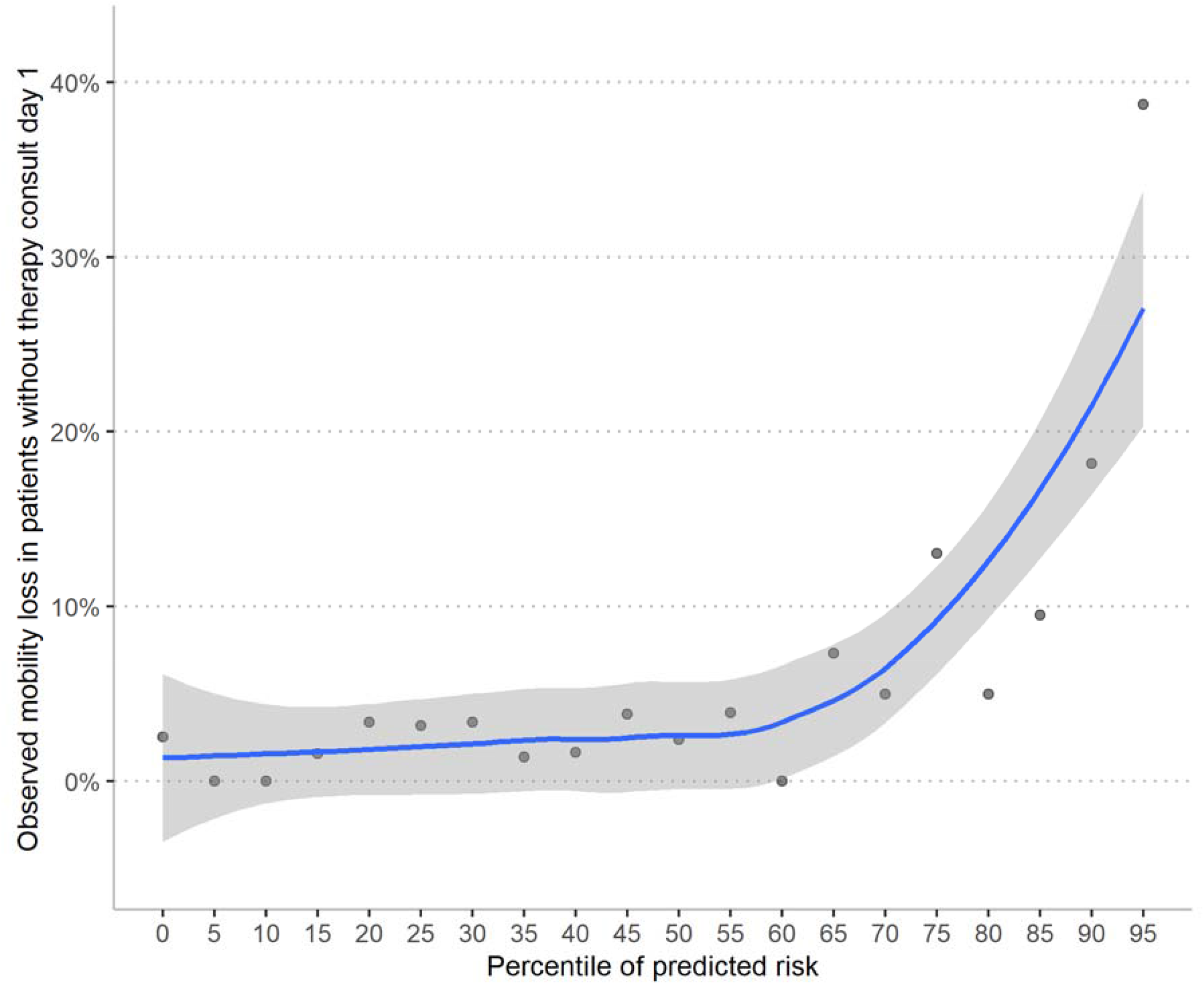
Mobility loss by predicted risk among those without a therapy consultation order on the first hospital day, validation cohort. <u>Legend</u> Analysis performed in the validation data among those who did not have a therapy consultation order on the first hospital day. Line fit using a loess regression with 90% span and weighted by number of observations in each ventile.

**Figure 3** illustrates how the model may identify patients at risk for mobility loss beyond clinical concern. **Figure 3** plots the rate of mobility loss by predicted risk in patients who did not have a therapy consult placed on the first hospital day. The loess regression demonstrates a curvilinear relationship where the rate of observed mobility loss increases exponentially after the 70^th^ percentile of predicted risk. For example, in the 95^th^ percentile group, among the 32% of patients who did not have a therapy consult on the first day, 39% were discharged with a new mobility impairment.

### Task-specific application

In the validation cohort, we describe test characteristics when the model results are dichotomized to support ‘rule out’ (i.e., high sensitivity) and ‘rule in’ (i.e., high specificity) tasks (**Table 3**). For a ‘rule out’ task, we dichotomized at a predicted risk of 2.7%, the 20^th^ percentile of predicted risk. At this threshold, the model produces a sensitivity of 97% (95% CI, 92% to 99%) and a specificity of 21% (95% CI, 20% to 23%). For a ‘rule in’ task, we dichotomized at a predicted risk of 11.0%, the 80^th^ percentile of predicted risk. At this threshold, the model produces a sensitivity of 49% (95% CI, 41% to 58%) and a specificity of 82% (95% CI, 81% to 84%).

**Table 3:** Predictive performance at 2 thresholds.

|  | **<u>Rule Out</u>**<br>**Predicted risk $\geq 2.7\%^*$**<br>**considered positive**<br>**(95% CI)** | **<u>Rule In</u>**<br>**Predicted risk $\geq 11.0\%^{**}$**<br>**considered positive**<br>**(95% CI)** |
| --- | --- | --- |
| PPV | 9.2% (8.9% to 9.5%) | 19% (16% to 22%) |
| NPV | 99% (97% to 100%) | 95% (94% to 96%) |
| Sensitivity | 97% (92% to 99%) | 49% (41% to 58%) |
| Specificity | 21% (20% to 23%) | 82% (81% to 84%) |

Test characteristics in the validation data with high sensitivity and high specificity thresholds. NPV, negative predicted value; PPV, positive predictive value
\* 20<sup>th</sup> percentile
\*\* 80<sup>th</sup> percentile

## DISCUSSION

In this study, we developed and validated the Day-1 Mobility Loss model to predict hospital-acquired mobility loss in older adults who were able to walk on admission. The model allows universal screening using routinely collected clinical and sociodemographic data on the first hospital day. The model performed well and demonstrated potential clinical application when tested in a validation cohort that was one year removed from the derivation cohort.

Several potential use cases exist for this rapid, universal screening model that uses routinely collected data in the electronic health record. First, in this study, the model identified patients with a greater-than-average risk of mobility loss who nevertheless did not have a therapy consultation ordered on the first hospital day. Targeting therapy consultations and mobility programs to this population may be particularly beneficial in staving off mobility loss. Targeting is salient because rehabilitation services and mobility programs are usually finite resources in hospitals. Beyond therapy consultations, identifying at-risk individuals could alert clinicians to the best practices for preventing hospital-acquired disability like avoiding bed rest orders, counseling patients to mobilize safely, limiting psychoactive medication use, reducing tether use, and attending to nutrition.^9^ Finally, because this model can be automated, it could be used as a “prescreen” for hospital-acquired disability models that are more accurate but require patient-reported data and therefore are more resource intensive to administer. For instance, this rapid, universal screen could identify intermediate and high-risk patients for screening with more accurate instruments that rely on cognitive assessments (e.g., Mini-Mental State Examination) and functional assessments (e.g., mobility 2 weeks before admission).^16–21^ A similar strategy used in screening for depressive symptoms where the Patient Health Questionnaire-2 (PHQ-2) used to prescreen patients for the more accurate yet resource-intensive PHQ-9.^30^

The study results also provided insight into physicians’ risk assessment of mobility loss. The study results indicated that while clinicians increased rates of therapy consultation ordering in those at increased risk, the increase was inadequate for those at highest risk. That is, we observed that ordering therapy consultations reflected an assumption that mobility loss risk increased linearly when, in fact, the risk increases exponentially. This finding redemonstrated exponential growth bias, a well-described cognitive bias described as the “pervasive tendency to linearize exponential functions when assessing them intuitively.”^31,32^ Properly implemented into clinical workflows, a prediction model like the one developed here can augment clinicians’ decision-making to mitigate this cognitive misestimation of risk.

Finally, this study adds to the literature on hospital-acquired mobility by identifying new predictors of hospital-acquired disability. Prior studies have identified age, cognition, prehospital functional status, delirium, and urinary catheters as substantive predictors of hospital-acquired disability.^9,33,34^ This study identifies medication administrations and mental status using Glasgow Coma Score as important predictors that have not been previously described.

The study design and data have limitations that are important to consider when interpreting the results. First, while this model identifies those at risk of losing the ability to walk during hospitalization, it should not be taken to mean that these are the only patients for whom mobility interventions should be used. Mobility interventions should also be used to restore patients to pre-illness baseline. For example, a patient who could walk independently but now walks with an assistive device may not be flagged by this model. Thus, this model should supplement, not replace clinician’s assessment of mobility loss risk. Second, the risk factors in the Day-1 Mobility Loss model are not necessarily causal; that is, it should not be taken to mean that addressing the risk factors in the model (e.g., removing a urinary catheter) identified will reduce mobility loss. Third, this model was developed and validated in a single academic center. The development and validation cohort were separated by a year, providing some assurance as to the generalizability of the model.^35^ Future external validation studies, particularly in community hospitals, will more completely define the generalizability of the Day-1 Mobility Loss model.

In conclusion, the Day-1 Mobility Loss model estimates the risk of mobility loss on the first hospital day in hospitalized older adults. The model uses data from the electronic medical record and does not require additional patient or clinician input. The model performed well and demonstrated the ability to meaningfully inform clinicians’ use of therapy resources.

## Data Availability

Data are not available due to UCSF restrictions on sharing of identifiable patient information.

## Author contributions

Dr. Shah had full access to all of the data in the study and takes responsibility for the integrity of the data and the accuracy of the data analysis. All authors listed have contributed sufficiently to the project to be included as authors, and all those who are qualified to be authors are listed in the author byline.

## Appendix 1: Braden activity subscale description

| Title (score) | Description |
| --- | --- |
| Bedfast (1) | Confined to bed |
| Chairfast (2) | Ability to walk severely limited or nonexistent |
| Walks occasionally (3) | Walks occasionally during the day but for very short distances, with or without assistance. Spends most each shift in the bed or chair |
| Walks frequently (4) | Walks outside of the room at least twice per day and inside of the room at least once every two hours during waking hours |

## Appendix 2: Cohort flow diagram

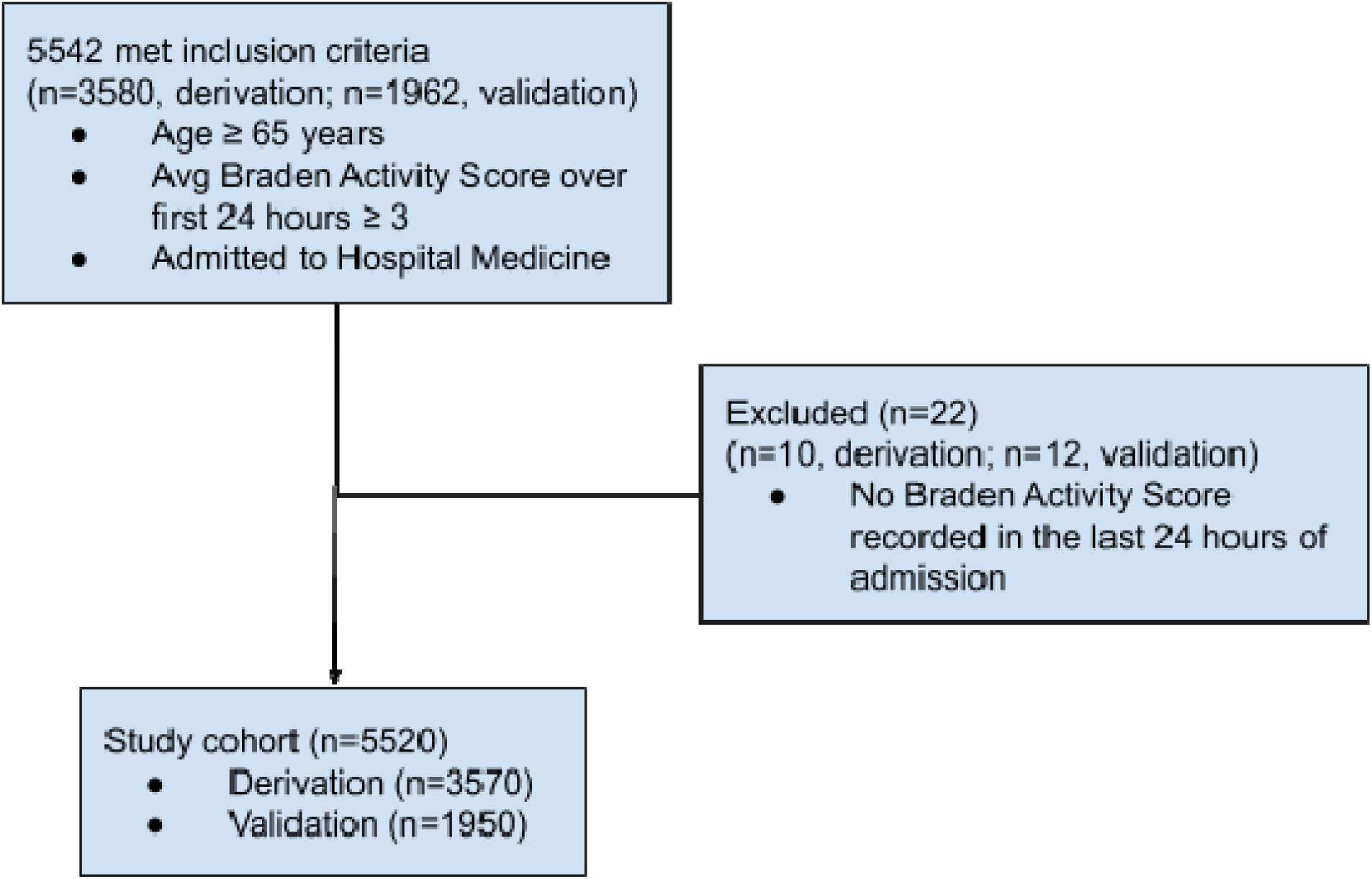

## Appendix 3: Functional form of continuous predictors

We considered various functional forms of the continuous candidate predictors. We fit the various functional forms as the only predictor of our outcome of interest in the derivation data set. We started with the linear functional form. If the BIC produced by an alternative functional form was one point lower than the BIC produced by the linear model, we used that functional form. Variables that did not produce a lower BIC than the intercept-only model were excluded altogether.

| Predictor | Intercept only | Linear | Log | Square | Categorical |
| --- | --- | --- | --- | --- | --- |
| Medication count | 1916* | 1922 | 1923 | 1923 | n/a |
| Medication administration | 1916 | 1858 | 1831* | 1907 | n/a |
| PIV count | 1916 | 1908* | 1911 | 1908 | n/a |
| Age | 1916 | 1906* | 1906 | 1906 | 1908 |
| GCS Verbal | 1916 | 1894 | 1900 | 1891 | 1886* |
| GCS Motor | 1916 | 1916 | 1917 | 1916 | 1915* |
| GCS Eyes | 1916 | 1913 | 1913 | 1913 | 1910* |
| Creatinine | 1916* | 1924 | 1920 | 1924 | 1922 |
| Albumin | 1916 | 1895 | 1893* | 1897 | 1905 |

## Appendix 4: Validation cohort calibration plot

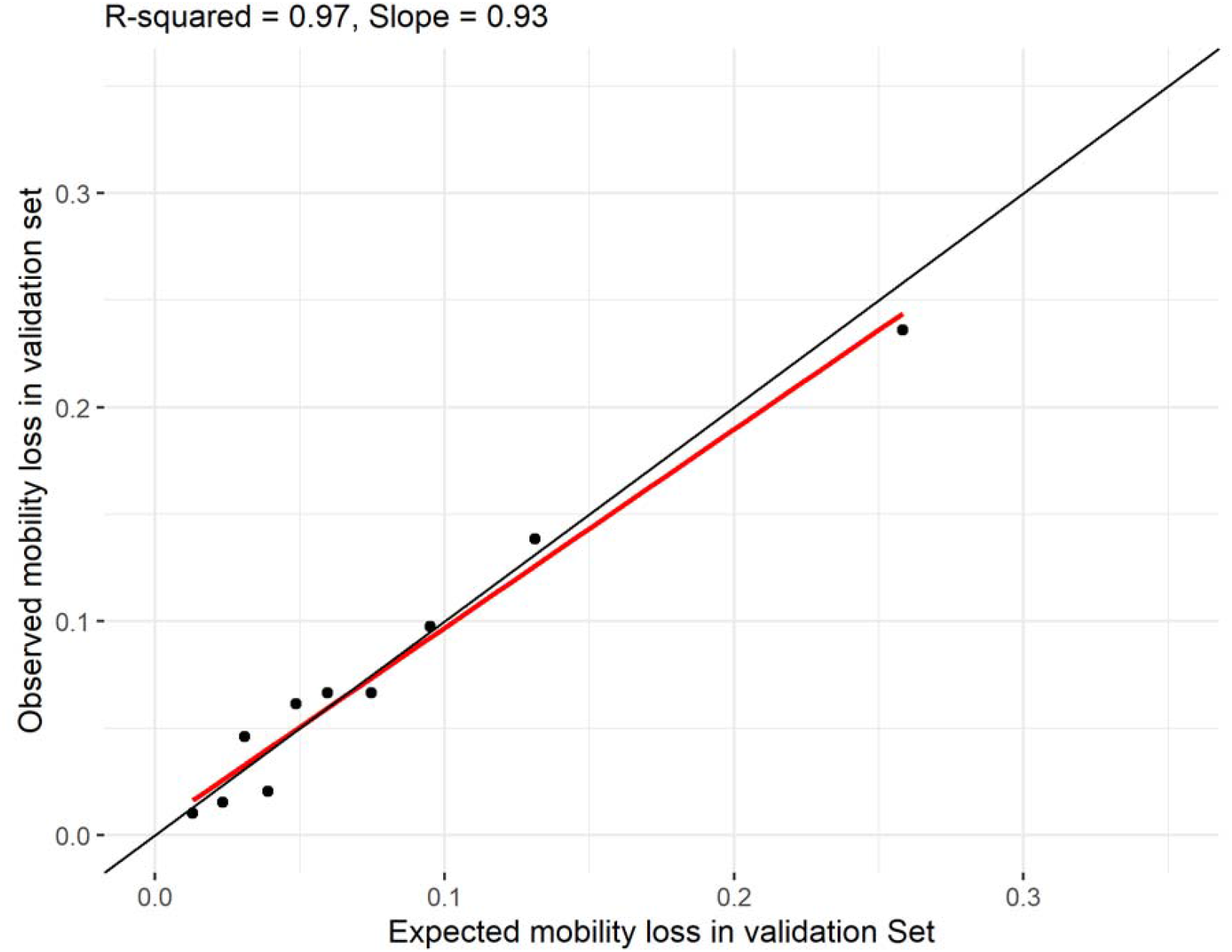

## Appendix 5: Receiver operator curve for mobility impairment prediction model in the development and validation cohort

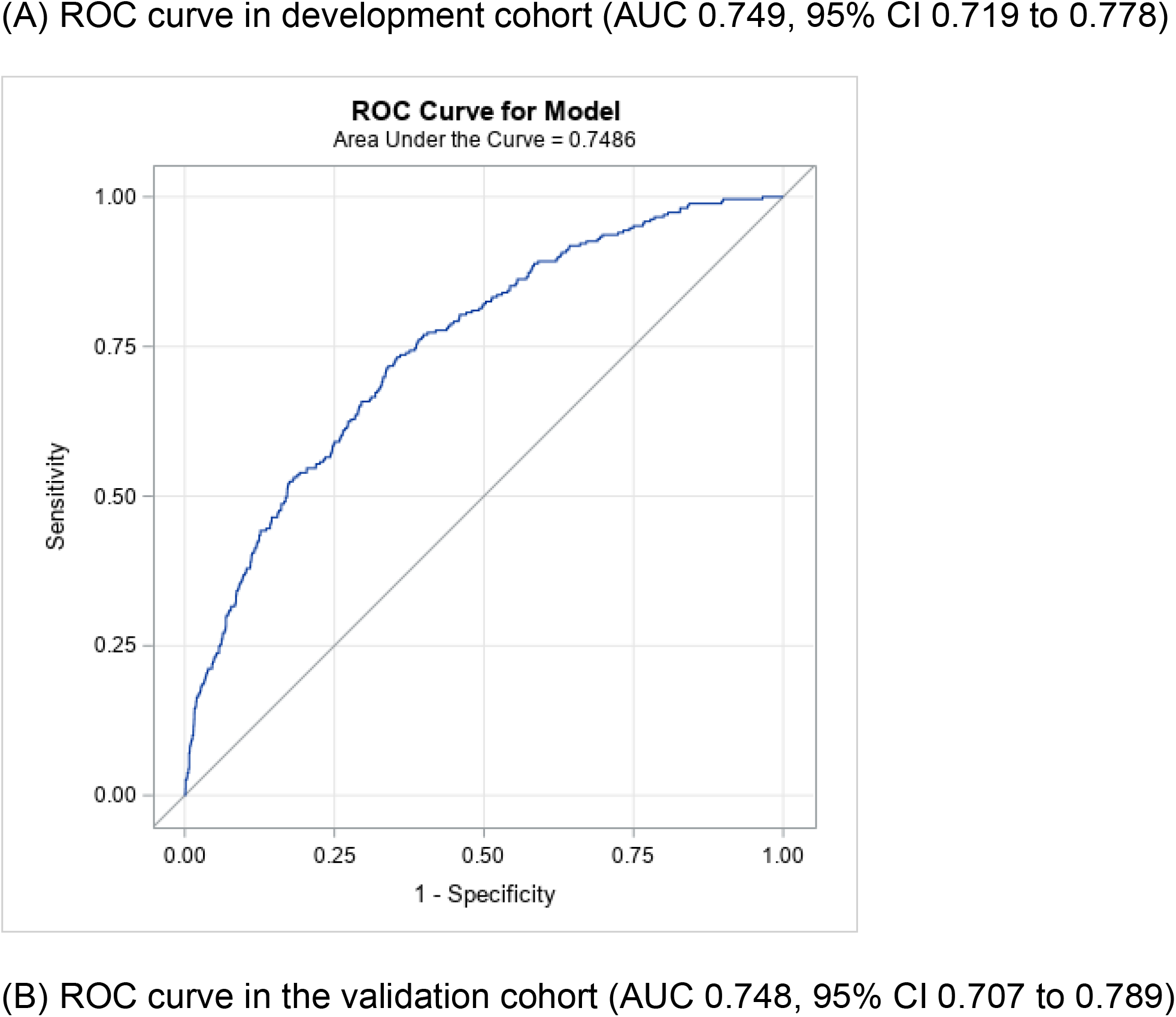
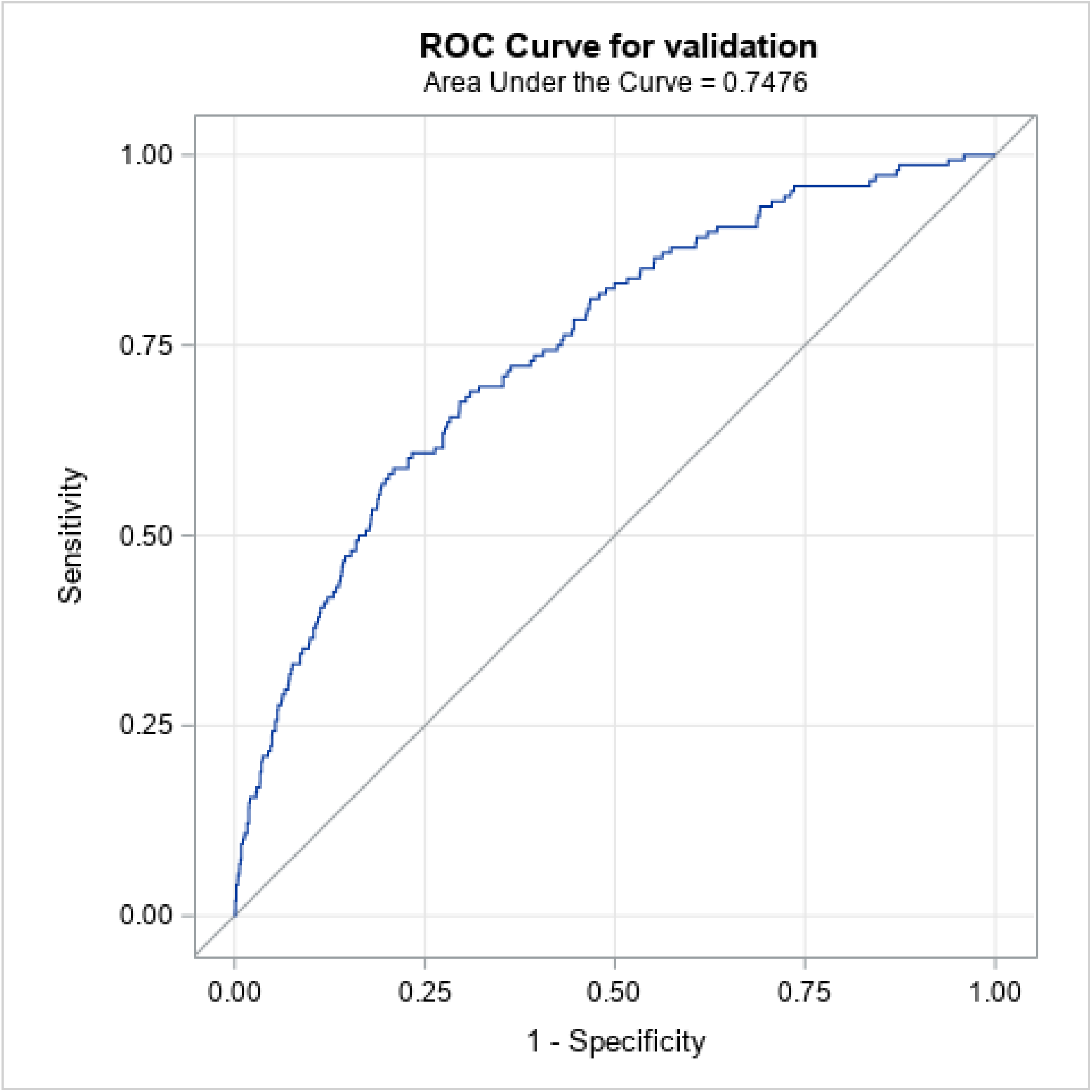

## Appendix 6: TRIPOD check list

| Section/Topic |  |  | Checklist Item | Page |
| --- | --- | --- | --- | --- |
| **Title and abstract** |  |  |  |  |
| Title | 1 | D;V | Identify the study as developing and/or validating a multivariable prediction model, the target population, and the outcome to be predicted. | 1 |
| Abstract | 2 | D;V | Provide a summary of objectives, study design, setting, participants, sample size, predictors, outcome, statistical analysis, results, and conclusions. | 2 |
| **Introduction** |  |  |  |  |
| Background and objectives | 3a | D;V | Explain the medical context (including whether diagnostic or prognostic) and rationale for developing or validating the multivariable prediction model, including references to existing models. | 3 |
|  | 3b | D;V | Specify the objectives, including whether the study describes the development or validation of the model or both. | 3 |
| **Methods** |  |  |  |  |
| Source of data | 4a | D;V | Describe the study design or source of data (e.g., randomized trial, cohort, or registry data), separately for the development and validation data sets, if applicable. | 4 |
|  | 4b | D;V | Specify the key study dates, including start of accrual; end of accrual; and, if applicable, end of follow-up. | 4 |
| Participants | 5a | D;V | Specify key elements of the study setting (e.g., primary care, secondary care, general population) including number and location of centres. | 4 |
|  | 5b | D;V | Describe eligibility criteria for participants. | 4 |
|  | 5c | D;V | Give details of treatments received, if relevant. | n/a |
| Outcome | 6a | D;V | Clearly define the outcome that is predicted by the prediction model, including how and when assessed. | 4 |
|  | 6b | D;V | Report any actions to blind assessment of the outcome to be predicted. | n/a |
| Predictors | 7a | D;V | Clearly define all predictors used in developing the multivariable prediction model, including how and when they were measured. | 4,5 |
|  | 7b | D;V | Report any actions to blind assessment of predictors for the outcome and other predictors. | n/a |
| Sample size | 8 | D;V | Explain how the study size was arrived at. | 4,<br>Appendix 1 |
| Missing data | 9 | D;V | Describe how missing data were handled (e.g., complete-case analysis, single imputation, multiple imputation) with details of any imputation method. | 5 |
| Statistical analysis methods | 10a | D | Describe how predictors were handled in the analyses. | 5 |
|  | 10b | D | Specify type of model, all model-building procedures (including any predictor selection), and method for internal validation. | 5 |
|  | 10c | V | For validation, describe how the predictions were calculated. | 5 |
|  | 10d | D;V | Specify all measures used to assess model performance and, if relevant, to compare multiple models. | 5 |
|  | 10e | V | Describe any model updating (e.g., recalibration) arising from the validation, if done. | n/a |
| Risk groups | 11 | D;V | Provide details on how risk groups were created, if done. | 5-6 |
| Development vs. validation | 12 | V | For validation, identify any differences from the development data in setting, eligibility criteria, outcome, and predictors. | 4 |
| **Results** |  |  |  |  |
| Participants | 13a | D;V | Describe the flow of participants through the study, including the number of participants with and without the outcome and, if applicable, a summary of the follow-up time. A diagram may be helpful. | Appendix 1 |
|  | 13b | D;V | Describe the characteristics of the participants (basic demographics, clinical features, available predictors), including the number of participants with missing data for predictors and outcome. | 6, Table 1 |
|  | 13c | V | For validation, show a comparison with the development data of the distribution of important variables (demographics, predictors and outcome). | Table 1 |
| Model development | 14a | D | Specify the number of participants and outcome events in each analysis. | 6 |
|  | 14b | D | If done, report the unadjusted association between each candidate predictor and outcome. | n/a |
| Model specification | 15a | D | Present the full prediction model to allow predictions for individuals (i.e., all regression coefficients, and model intercept or baseline survival at a given time point). | Table 2 |
|  | 15b | D | Explain how to use the prediction model. | Table 2 |
| Model performance | 16 | D;V | Report performance measures (with CIs) for the prediction model. | Table 3 |
| Model-updating | 17 | V | If done, report the results from any model updating (i.e., model specification, model performance). | n/a |
| **Discussion** |  |  |  |  |
| Limitations | 18 | D;V | Discuss any limitations of the study (such as nonrepresentative sample, few events per predictor, missing data). | 9, 10 |
| Interpretation | 19a | V | For validation, discuss the results with reference to performance in the development data, and any other validation data. | n/a |
|  | 19b | D;V | Give an overall interpretation of the results, considering objectives, limitations, results from similar studies, and other relevant evidence. | 8-10 |
| Implications | 20 | D;V | Discuss the potential clinical use of the model and implications for future research. | 9-10 |
| **Other information** |  |  |  |  |
| Supplementary information | 21 | D;V | Provide information about the availability of supplementary resources, such as study protocol, Web calculator, and data sets. | N/a |
| Funding | 22 | D;V | Give the source of funding and the role of the funders for the present study. | 11 |

\*Items relevant only to the development of a prediction model are denoted by D, items relating solely to a validation of a prediction model are denoted by V, and items relating to both are denoted D;V. We recommend using the TRIPOD Checklist in conjunction with the TRIPOD Explanation and Elaboration document.

## References

1. Hoyer EH, Needham DM, Atanelov L, Knox B, Friedman M, Brotman DJ. Association of impaired functional status at hospital discharge and subsequent rehospitalization. J Hosp Med. 2014;9(5):277–282. doi:10.1002/jhm.2152

2. Lang PO, Heitz D, Hédelin G, et al. Early markers of prolonged hospital stays in older people: a prospective, multicenter study of 908 inpatients in French acute hospitals. J Am Geriatr Soc. 2006;54(7):1031–1039. doi:10.1111/j.1532-5415.2006.00767.x

3. Brown CJ, Friedkin RJ, Inouye SK. Prevalence and outcomes of low mobility in hospitalized older patients. J Am Geriatr Soc. 2004;52(8):1263–1270. doi:10.1111/j.1532-5415.2004.52354.x

4. Brown CJ, Flood KL. Mobility limitation in the older patient: a clinical review. JAMA. 2013;310(11):1168–1177. doi:10.1001/jama.2013.276566

5. Agmon M, Zisberg A, Gil E, Rand D, Gur-Yaish N, Azriel M. Association Between 900 Steps a Day and Functional Decline in Older Hospitalized Patients. JAMA Intern Med. 2017;177(2):272. doi:10.1001/jamainternmed.2016.7266

6. Pedersen MM, Bodilsen AC, Petersen J, et al. Twenty-Four-Hour Mobility During Acute Hospitalization in Older Medical Patients. The Journals of Gerontology Series A: Biological Sciences and Medical Sciences. 2013;68(3):331–337. doi:10.1093/gerona/gls165

7. Brown CJ, Redden DT, Flood KL, Allman RM. The underrecognized epidemic of low mobility during hospitalization of older adults. J Am Geriatr Soc. 2009;57(9):1660–1665. doi:10.1111/j.1532-5415.2009.02393.x

8. Covinsky KE, Palmer RM, Fortinsky RH, et al. Loss of Independence in Activities of Daily Living in Older Adults Hospitalized with Medical Illnesses: Increased Vulnerability with Age. Journal of the American Geriatrics Society. 2003;51(4):451–458. doi:10.1046/j.1532-5415.2003.51152.x

9. Covinsky KE, Pierluissi E, Johnston CB. Hospitalization-Associated Disability: “She Was Probably Able to Ambulate, but I’m Not Sure.” JAMA. 2011;306(16):1782–1793. doi:10.1001/jama.2011.1556

10. Boyd CM, Landefeld CS, Counsell SR, et al. Recovery of Activities of Daily Living in Older Adults After Hospitalization for Acute Medical Illness. Journal of the American Geriatrics Society. 2008;56(12):2171–2179. doi:10.1111/j.1532-5415.2008.02023.x

11. Dharmarajan K, Han L, Gahbauer EA, Leo-Summers LS, Gill TM. Disability and Recovery After Hospitalization for Medical Illness Among Community-Living Older Persons: A Prospective Cohort Study. Journal of the American Geriatrics Society. 2020;68(3):486–495. doi:10.1111/jgs.16350

12. Adler J, Malone D. Early mobilization in the intensive care unit: a systematic review. Cardiopulm Phys Ther J. 2012;23(1):5–13.

13. Hoyer EH, Friedman M, Lavezza A, et al. Promoting mobility and reducing length of stay in hospitalized general medicine patients: A quality-improvement project. J Hosp Med. 2016;11(5):341–347. doi:10.1002/jhm.2546

14. den Hertog A, Gliesche K, Timm J, Mühlbauer B, Zebrowski S. Pathway-controlled fast-track rehabilitation after total knee arthroplasty: a randomized prospective clinical study evaluating the recovery pattern, drug consumption, and length of stay. Arch Orthop Trauma Surg. 2012;132(8):1153–1163. doi:10.1007/s00402-012-1528-1

15. Cohen Y, Zisberg A, Chayat Y, et al. Walking for Better Outcomes and Recovery: The Effect of WALK-FOR in Preventing Hospital-Associated Functional Decline Among Older Adults. Newman A, ed. The Journals of Gerontology: Series A. 2019;74(10):1664–1670. doi:10.1093/gerona/glz025

16. Inouye SK, Wagner DR, Acampora D, et al. A predictive index for functional decline in hospitalized elderly medical patients. J Gen Intern Med. 1993;8(12):645–652. doi:10.1007/BF02598279

17. Mehta KM, Pierluissi E, Boscardin WJ, et al. A Clinical Index to Stratify Hospitalized Older Adults According to Risk for New-Onset Disability. Journal of the American Geriatrics Society. 2011;59(7):1206–1216. doi:10.1111/j.1532-5415.2011.03409.x

18. Sager MA, Rudberg MA, Jalaluddin M, et al. Hospital Admission Risk Profile (HARP): Identifying Older Patients at Risk for Functional Decline Following Acute Medical Illness and Hospitalization. Journal of the American Geriatrics Society. 1996;44(3):251–257. doi:10.1111/j.1532-5415.1996.tb00910.x

19. Huyse FJ, de Jonge P, Slaets JPJ, et al. COMPRI—An Instrument to Detect Patients With Complex Care Needs: Results From a European Study. Psychosomatics. 2001;42(3):222–228. doi:10.1176/appi.psy.42.3.222

20. McCusker J, Bellavance F, Cardin S, Trepanier S, Verdon J, Ardman O. Detection of Older People at Increased Risk of Adverse Health Outcomes After an Emergency Visit: The ISAR Screening Tool. Journal of the American Geriatrics Society. 1999;47(10):1229–1237. doi:10.1111/j.1532-5415.1999.tb05204.x

21. Geyskens L, Jeuris A, Deschodt M, Van Grootven B, Gielen E, Flamaing J. Patient-related risk factors for in-hospital functional decline in older adults: A systematic review and meta-analysis. Age and Ageing. 2022;51(2):afac007. doi:10.1093/ageing/afac007

22. Bergstrom N, Braden BJ, Laguzza A, Holman V. The Braden Scale for Predicting Pressure Sore Risk. Nurs Res. 1987;36(4):205–210.

23. Valiani V, Chen Z, Lipori G, Pahor M, Sabbá C, Manini TM. Prognostic Value of Braden Activity Subscale for Mobility Status in Hospitalized Older Adults. J Hosp Med. 2017;12(6):396–401. doi:10.12788/jhm.2748

24. Chase JAD, Lozano A, Hanlon A, Bowles KH. Identifying Factors Associated With Mobility Decline Among Hospitalized Older Adults. Clin Nurs Res. 2018;27(1):81–104. doi:10.1177/1054773816677063

25. Teasdale G, Maas A, Lecky F, Manley G, Stocchetti N, Murray G. The Glasgow Coma Scale at 40 years: standing the test of time. The Lancet Neurology. 2014;13(8):844–854. doi:10.1016/S1474-4422(14)70120-6

26. Teasdale G, Jennett B. ASSESSMENT OF COMA AND IMPAIRED CONSCIOUSNESS: A Practical Scale. The Lancet. 1974;304(7872):81–84. doi:10.1016/S0140-6736(74)91639-0

27. Steyerberg EW, Vickers AJ, Cook NR, et al. Assessing the performance of prediction models: a framework for some traditional and novel measures. Epidemiology. 2010;21(1):128–138. doi:10.1097/EDE.0b013e3181c30fb2

28. Oseran AS, Lage DE, Jernigan MC, Metlay JP, Shah SJ. A “Hospital-Day-1” Model to Predict the Risk of Discharge to a Skilled Nursing Facility. Journal of the American Medical Directors Association. 2019;20(6):689-695.e5. doi:10.1016/j.jamda.2019.03.035

29. Singer DE, Chang Y, Borowsky LH, et al. A New Risk Scheme to Predict Ischemic Stroke and Other Thromboembolism in Atrial Fibrillation: The ATRIA Study Stroke Risk Score. J Am Heart Assoc. 2013;2(3):e000250. doi:10.1161/JAHA.113.000250

30. Levis B, Sun Y, He C, et al. Accuracy of the PHQ-2 Alone and in Combination With the PHQ-9 for Screening to Detect Major Depression: Systematic Review and Meta-analysis. JAMA. 2020;323(22):2290. doi:10.1001/jama.2020.6504

31. Banerjee R, Bhattacharya J, Majumdar P. Exponential-growth prediction bias and compliance with safety measures related to COVID-19. Social Science & Medicine. 2021;268:113473. doi:10.1016/j.socscimed.2020.113473

32. Stango V, Zinman J. Exponential Growth Bias and Household Finance. The Journal of Finance. 2009;64(6):2807–2849. doi:10.1111/j.1540-6261.2009.01518.x

33. Hoogerduijn JG, Schuurmans MJ, Duijnstee MS, Rooij SED, Grypdonck MF. A systematic review of predictors and screening instruments to identify older hospitalized patients at risk for functional decline. Journal of Clinical Nursing. 2007;16(1):46–57. doi:10.1111/j.1365-2702.2006.01579.x

34. Zisberg A, Shadmi E, Gur-Yaish N, Tonkikh O, Sinoff G. Hospital-Associated Functional Decline: The Role of Hospitalization Processes Beyond Individual Risk Factors. Journal of the American Geriatrics Society. 2015;63(1):55–62. doi:10.1111/jgs.13193

35. Justice AC, Covinsky KE, Berlin JA. Assessing the Generalizability of Prognostic Information. Ann Intern Med. 1999;130(6):515–524. doi:10.7326/0003-4819-130-6-199903160-00016

